# Lipid storm within the lungs of severe COVID-19 patients: Extensive levels of cyclooxygenase and lipoxygenase-derived inflammatory metabolites

**DOI:** 10.1101/2020.12.04.20242115

**Authors:** Anne-Sophie Archambault, Younes Zaid, Volatiana Rakotoarivelo, Étienne Doré, Isabelle Dubuc, Cyril Martin, Youssef Amar, Amine Cheikh, Hakima Fares, Amine El Hassani, Youssef Tijani, Michel Laviolette, Éric Boilard, Louis Flamand, Nicolas Flamand

## Abstract

**BACKGROUND:** Severe Acute Respiratory Syndrome coronavirus 2 (SARS-CoV-2) is the infectious agent responsible for Coronavirus disease 2019 (COVID-19). While SARS-CoV-2 infections are often benign, there are also severe COVID-19 cases, characterized by severe bilobar pneumonia that can decompensate to an acute respiratory distress syndrome, notably characterized by increased inflammation and a cytokine storm. While there is no cure against severe COVID-19 cases, some treatments significantly decrease the severity of the disease, notably aspirin and dexamethasone, which both directly or indirectly target the biosynthesis (and effects) of numerous bioactive lipids.

**OBJECTIVE:** Our working hypothesis was that severe COVID-19 cases necessitating mechanical ventilation were characterized by increased bioactive lipid levels modulating lung inflammation. We thus quantitated several lung bioactive lipids using liquid chromatography combined to tandem mass spectrometry.

**RESULTS:** We performed an exhaustive assessment of the lipid content of bronchoalveolar lavages from 25 healthy controls and 33 COVID-19 patients necessitating mechanical ventilation. Severe COVID-19 patients were characterized by increased fatty acid levels as well as an accompanying inflammatory lipid storm. As such, most quantified bioactive lipids were heavily increased. There was a predominance of cyclooxygenase metabolites, notably TXB_2_ >> PGE_2_ ∼ 12-HHTrE > PGD_2_. Leukotrienes were also increased, notably LTB_4_, 20-COOH-LTB_4_, LTE_4_, and eoxin E_4_. 15-lipoxygenase metabolites derived from linoleic, arachidonic, eicosapentaenoic and docosahexaenoic acids were also increased. Finally, yet importantly, specialized pro-resolving mediators, notably lipoxin A_4_ and the D-series resolvins, were also found at important levels, underscoring that the lipid storm occurring in severe SARS-CoV-2 infections involves pro- and anti-inflammatory lipids.

**CONCLUSIONS:** Our data unmask the important lipid storm occurring in the lungs of patients afflicted with severe COVID-19. We discuss which clinically available drugs could be helpful at modulating the lipidome we observed in the hope of minimizing the deleterious effects of pro-inflammatory lipids and enhancing the effects of anti-inflammatory and/or pro-resolving lipids.

## INTRODUCTION

Coronavirus disease 2019 (COVID-19) is an infectious disease caused by the Severe Acute Respiratory Syndrome coronavirus 2 (SARS-CoV-2) (1). The clinical manifestations of the disease are often mild but severe cases can lead to bilobar pneumonia with hyperactivation of the inflammatory cascade and progression to acute respiratory distress syndrome (ARDS), necessitating mechanical ventilation (2, 3). SARS-CoV-2 spreads predominantly from respiratory droplets of infected individuals to mucosal epithelial cells in the upper airways and oral cavity (4). It infects cells *via* its homotrimeric spike protein binding to host-cell expressing angiotensin-converting enzyme-2 (ACE2) receptor in a protease-dependent manner (5).

Autopsies from severe COVID-19 patients unmasked a heterogeneous disease that can be characterized by diffuse alveolar damage, endothelial damage, thrombosis of small and medium vessels, pulmonary embolism, and inflammatory cell infiltration, sometimes more lymphocytic, sometimes more granulocytic (3, 6-8). An uncontrolled systemic inflammatory response known as the cytokine storm also occurs. This cytokine storm, which is the consequence of an important release of pro-inflammatory cytokines, such as tumor necrosis factor-α, interleukin (IL)-1β, IL-6, IL-7 and granulocyte-colony stimulating factor, is suggested as an important contributor of SARS-CoV-2 lethality (8, 9). However, the heterogeneity of the disease indicates that, aside cytokines (and chemokines), other immunological effectors contribute to the sustained inflammatory responses observed in the most severe forms of COVID-19.

Among the numerous soluble effectors involved in the infectious/inflammatory response, bioactive lipids such as eicosanoids likely promote their fair share of deleterious effects, notably by enhancing leukocyte recruitment and activation, by participating in the exudate formation and by stimulating platelet aggregation and thrombus formation. In contrast, specialized pro-resolving mediators (SPMs), which mainly consist of docosanoids, could dampen the inflammatory response and even promote its resolution (10). While therapeutic approaches targeting the biosynthesis or effects of inflammatory lipids are readily available, the levels of those lipids in the lungs during severe COVID-19 remain unknown.

Herein, we performed a detailed lipidomic analysis of bronchoalveolar lavage (BAL) fluids from healthy volunteers and severe COVID-19 patients. We provide evidence that severe COVID-19 patients are characterized by robust levels of lipids arising from the cyclooxygenase (COX) and lipoxygenase (LOX) pathways as well increased SPM levels. Our study further highlights that few associations exist between clinical parameters and bioactive lipids present in the primary site of disease, the lungs.

## MATERIAL AND METHODS

### Materials

All lipids were obtained from Cayman Chemical (Ann Arbor, MI, USA) unless indicated otherwise. DMSO and C17:1-LPA were purchased from Sigma-Aldrich (St-Louis, MO, USA). DMSO, ammonium acetate, acetic acid, LC-MS-grade MeOH, MeCN and H_2_O were purchased from Fisher Scientific (Ottawa, Canada). Cytokine/chemokine bead arrays were purchased from Becton Dickinson (BD).

### Ethics

This study was approved by the Local Ethics Committee of Cheikh Zaid Hospital (Project: CEFCZ/PR/2020-PR04), Rabat, Morocco and complies with the Declaration of Helsinki and all subjects signed a consent form. The analysis of bioactive lipids in the BAL of healthy donors and severe COVID-19 patients was approved by the Comité d’éthique de la recherche de l’Institut universitaire de cardiologie et de pneumologie de Québec – Université Laval and the Comité d’éthique de la recherche du CHU de Québec-Université Laval.

#### Clinical criteria for subject selection

Non-smoking healthy subjects taking no other medication than anovulants, without any documented acute or chronic inflammatory disease and without recent (8 weeks) airway infection were recruited for bronchoalveolar lavages at Institut universitaire de cardiologie et pneumologie de Québec (Québec City, Canada). Only BALs in which the cell content in alveolar macrophages was greater than 95%, with lymphocytes as the other main cell type, were kept for further analyses. Severe COVID-19 patients were enrolled based on the inclusion criteria for intubation and the need of mechanical ventilation support at Cheikh Zaid Hospital (Rabat, Morocco). Following intubation and the initiation of mechanical ventilation, blood sampling was done to assess the clinical variables shown in Table 1. Patients were ventilated in the prone decubitus position with a tidal volume of 6 ml/kg and a PEEP varying between 8 – 14 cm H_2_O. The FiO_2_ was adjusted between 0.6 – 1.0 to obtain a SpO_2_ ≥ 92%.

**Table 1.** Clinical characteristics of BAL donors.

|  | **Severe COVID-19** | **Healthy controls** |
| --- | --- | --- |
| **N** | 33 | 25 |
| **Women/Men** | 16/17 | 16/9 |
| **Death (Y/N)** | 2/31 | N/A |
| **SARS-CoV-2 detection in BAL (Ct)** | 25.303 ± 0.906 | N/A |
| **Age** | 58.455 ± 3.175 | 26.130 ± 1.030 |
| **Weight (kg)** | 73.097 ± 2.873 | N/A |
| **Hospital stay (days)** | 20.545 ± 2.014 | N/A |
| **Days before BAL** | 3.485 ± 0.763 | N/A |
| **D-Dimers (mg/ml)** | 1.210 ± 0.129 | N/A |
| **CRP (mg/l)** | 20.652 ± 2.525 | N/A |
| **ALT (U/l)** | 32.582 ± 2.144 | N/A |
| **AST (U/l)** | 35.145 ± 2.978 | N/A |
| **LDH (U/l)** | 616.152 ± 41.175 | N/A |
| **Hemoglobin (g/l)** | 123.152 ± 2.365 | N/A |
| **Blood Monocytes (million/ml)** | 0.327 ± 0.034 | N/A |
| **Blood Neutrophils (million/ml)** | 2.855 ± 0.194 | N/A |
| **Blood Eosinophils (million/ml)** | 0.026 ± 0.003 | N/A |
| **Blood Platelets (million/ml)** | 181.848 ± 13.397 | N/A |
| **Blood Lymphocytes (million/ml)** | 1.127 ± 0.127 | N/A |
| **Platelet to Lymphocyte ratio** | 223.893 ± 34.934 | N/A |
| **BAL Neutrophils (million/ml)** | 24.694 ± 2.152 | N/A |
| **BAL Eosinophils (million/ml)** | 0.197 ± 0.077 | N/A |
| **BAL Lymphocytes (million/ml)** | 21.830 ± 2.393 | N/A |

Data are presented as the mean ± SEM. **ALT**, alanine transaminase; **AST**, aspartate transaminase; **CRP**, C-reactive protein; **LDH**, Lactate dehydrogenase; **N/A**, not available;

#### Bronchoalveolar lavages

BAL fluids from healthy volunteers were obtained as follows: One 50 ml bolus of sterile 0.9% saline was injected in a sub-segmental bronchi of the right middle lobe. The obtained lavages were centrifuged (350 × *g*, 10 minutes) to pellet cells and the supernatants were immediately frozen (−80° Celsius) until further processing. A 5 ml aliquot of the samples was thawed then reduced to ∼500 µl using a stream of nitrogen. BAL fluids from severe COVID-19 patients were obtained less than 2 hrs following intubation and as follows: a total of 100 ml of sterile 0.9% saline (2 boli of 50 ml) was injected in a sub-segmental bronchi of the right middle lobe. The obtained lavages were pooled and centrifuged (320 × *g*, 15 minutes) to pellet cells. Supernatants were next concentrated as for the BALs of healthy donors using a stream of nitrogen and immediately frozen (−80° Celsius) until further processing. All of the evaporated samples were processed for LC-MS/MS analysis as described below.

#### LC-MS/MS analyses

Samples (500 µl) were denatured by adding 500 µl of LC-MS grade MeOH containing the internal standards then warmed at 60° Celsius for 30 minutes to inactivate (or not) SARS-CoV-2. Samples were then denatured overnight (−20° Celsius). The morning after, samples were warmed to room temperature and centrifuged at 10,000 × *g* to remove the denaturated proteins. The obtained supernatants were diluted with water containing 0.01% acetic acid to obtain a MeOH concentration of 10%, then acidified to pH 3 with acetic acid. Lipids were extracted by solid phase extraction (SPE) using Strata-X cartridges (Polymeric Reversed Phase, 60 mg/1 ml, Phenomenex, Torrance, CA, USA) as described before (11). In brief, columns were preconditioned with 2 ml MeOH containing 0.01% acetic acid followed by 2 ml LC-MS grade H_2_O containing 0.01% acetic acid. After sample loading, the columns were washed with 2 ml LC-MS grade H_2_O containing 0.01% acetic acid. Lipids were then eluted from the cartridges with 1 ml MeOH containing 0.01% acetic acid. Eluates were dried under a stream of nitrogen and reconstituted with 25 µl of solvent A (H_2_O + 1 mM NH_4_ + 0.05% acetic acid) and 25 µl of solvent B (MeCN/H_2_O, 95/5, v/v + 1 mM NH_4_ + 0.05% acetic acid). A volume of 40 µl of the resulting mixture was injected onto a RP-HPLC column (Kinetex C8, 150 × 2.1 mm, 2.6 µm, Phenomenex). Samples were eluted at a flow rate of 400 µl/min with a linear gradient of 10% solvent B that increased to 35% in 2 min, up to 75% in 10 min, from 75% to 95% in 0.1 min, and held at 98% for 5 min before re-equilibration to 10% solvent B for 2 min. The HPLC system was directly interfaced into the electrospray source of a Shimadzu 8050 triple quadrupole mass spectrometer and mass spectrometric analyses were performed in the positive (+) or the negative (−) ion mode using multiple reaction monitoring for the specific mass transitions of each lipid (Table 2). Quantification of each compound was done using internal standards and calibration curves that were extracted by solid-phase extraction as described above.

**Table 2.**
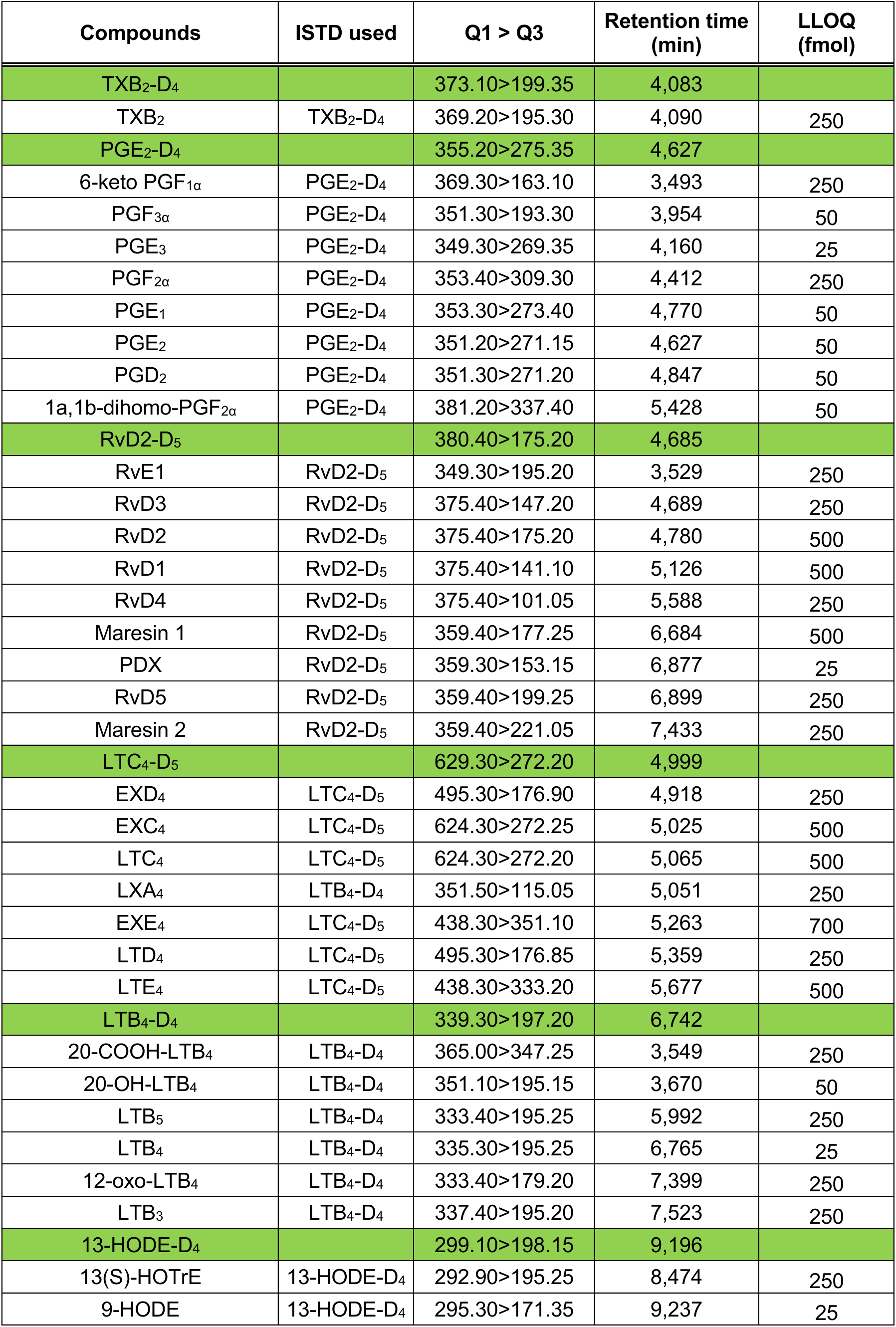
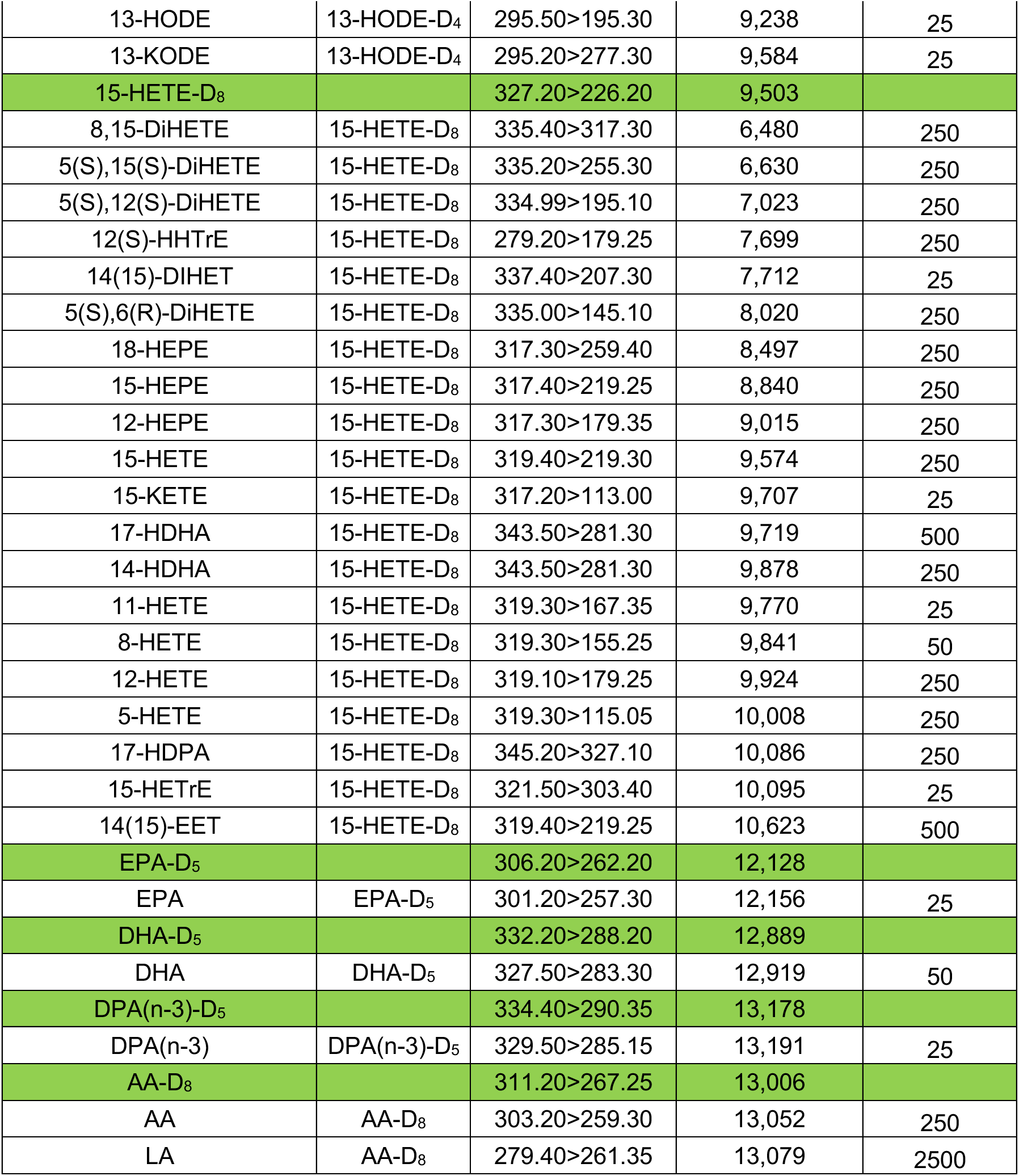
Mass transition of the investigated lipid mediators.

### Statistical analysis

Statistical analyses were done using the GraphPad Prism 9 software. P values < 0.05 were considered significant.

## RESULTS

While the circulating (plasmatic) cytokines’ profile observed in severe COVID-19 patients is relatively well-defined (8, 12-14), a much more limited number of studies characterizing the cytokine storm in the lungs of severe COVID-19 patients were available (13, 15, 16). In fact, to the best of our knowledge, no study simultaneously quantitated numerous soluble mediators in BAL fluids. We thus performed their lipidomic analysis in order to define whether an inflammatory lipid storm was present in the lungs of severe COVID-19 patients. Samples were compared to those obtained from healthy subjects. As a starting point, we investigated eicosanoids derived from the cyclooxygenase (COX) pathway. Our working hypothesis was that some COX metabolites would be increased, considering that IL-1β levels are elevated in the BALs of some COVID-19 patients (16) and that IL-1β is a potent inducer of cyclooxygenase (COX)-2 expression (17-19). The only COX metabolites that were detected in the BALs of healthy subjects were PGD_2_ (16 donors out of 25) and 12-HHTrE (2 donors out of 25). In contrast, BALs from severe COVID-19 patients had a significant increase of all COX metabolites investigated (**figure 1**). Most samples had detectable levels of PGD_2_ (27 of 33 donors) and the PGI_2_ metabolite 6-keto-PGF_1α_ (16 of 33 donors), while all samples contained PGF_2α_, important levels of PGE_2_, the inactive TXA_2_ metabolite TXB_2_, and the thromboxane synthase metabolite 12-HHTrE (**figure 1A**). Although there was a significant increase in all metabolites, the most increased COX-derived metabolites were TXB_2_ >> PGE_2_ ∼ 12-HHTrE > PGD_2_. We next addressed if the different COX metabolites correlated with each other. The levels of TXB_2_ strongly correlated with those of 12-HHTrE, PGD_2_, PGE_2_ and PGF_2α_ (**figure 1B-E**). Of note, few correlations were found between COX metabolites and the clinical parameters shown in Table1. To that end, PGF_2α_ and 12-HHTrE weakly but significantly correlated negatively with AST (**figure 7**),

**Figure 1.**
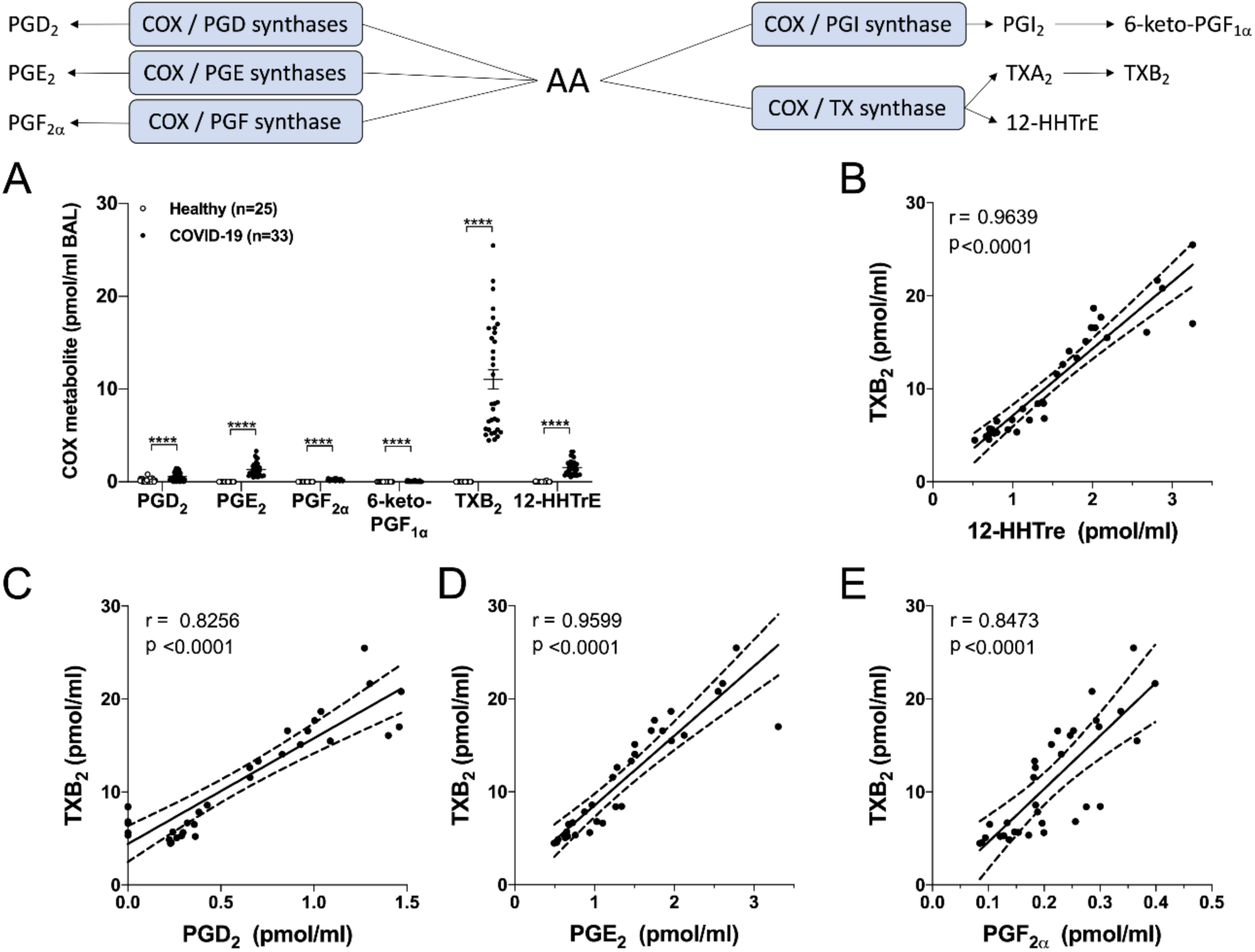
Levels of cyclooxygenase-derived metabolites in the BAL fluids of healthy and severe COVID-19 patients. BALs were obtained and processed as described in *Methods*. **A)** Results are from the BALs from 25 healthy subjects and 33 severe COVID-19 patients. P values were obtained by performing a Mann-Whitney test: **** = p < 0.0001. **B-E)** Correlation tests were performed with COVID-19 patients by using the non-parametric Spearman’s rank. P<0.05 was the threshold of significance. The linear regression is represented with a solid line and the best-fit lines of 95% confidence are represented with dotted lines.

Leukotrienes are also potent bioactive lipids previously documented to regulate the inflammatory responses in the airway/lung in diseases, notably ARDS (20, 21). As such, their involvement in the pathogenesis of COVID-19 has recently been postulated even though their levels were unknown (22). We thus quantitated the levels of leukotrienes derived from the 5-lipoxygenase pathway as well as those derived from the 15-lipoxygenase pathway, the 14,15-leukotrienes, which are now classified as eoxins (23). Leukotrienes were not detected in the BAL fluids of healthy volunteers, with the exception of LTB_4_, which was detected in 4 of 25 healthy subjects. In contrast, COVID-19 patients were characterized by increased levels of LTB_4_ and its CYP4F3A metabolite 20-COOH-LTB_4_ (**figure 2A**) as well as other minor metabolites (20-OH-LTB_4_ and 12-oxo-LTB_4_). Of note, the levels of LTB_4_ found in the BAL fluids of severe COVID-19 patients are comparable to those found in BAL fluids of ARDS patients (16). As expected, there was a strong correlation between the levels of LTB_4_ and those of 20-COOH-LTB_4_ (**figure 2B**). Importantly, the ratio between the sum of 20-OH- and 20-COOH-LTB_4_ to LTB_4_ ((20-OH- + 20- COOH-LTB_4_)/LTB_4_) was 2.843 +/- 0.532 (mean +/- SEM), indicating an increased omega oxidation of LTB_4_. The levels of LTB_4_ strongly correlated with those of TXB_2_ (**figure 2C**). The cysteinyl-leukotrienes LTE_4_ and EXE_4_ were also found in most severe COVID-19 patients (**figure 2D**). Surprisingly, the levels of EXE_4_ trended to be higher than those of LTE_4_. Furthermore, there was a trend toward a negative correlation between LTE_4_ and EXE_4_ levels although it did not reach statistical significance (**figure 2E**). This suggests that some COVID-19 patients are more prone to biosynthesize the 15-lipoxygnease-derived EXC_4_ while others are more prone to biosynthesize the 5-lipoxygenase-derived LTC_4_. Of note, LTB_4_ levels correlated with those of EXE_4_ but not with those of LTE_4_ (**figure 2F,G**). Again, a limited number of correlations between the different leukotrienes and the clinical parameters from Table 1 were found (**figure 7**), supporting the concept that the levels of leukotrienes in the lungs are not reflected in the blood by the most recognized COVID-19-related biomarkers.

**Figure 2.**
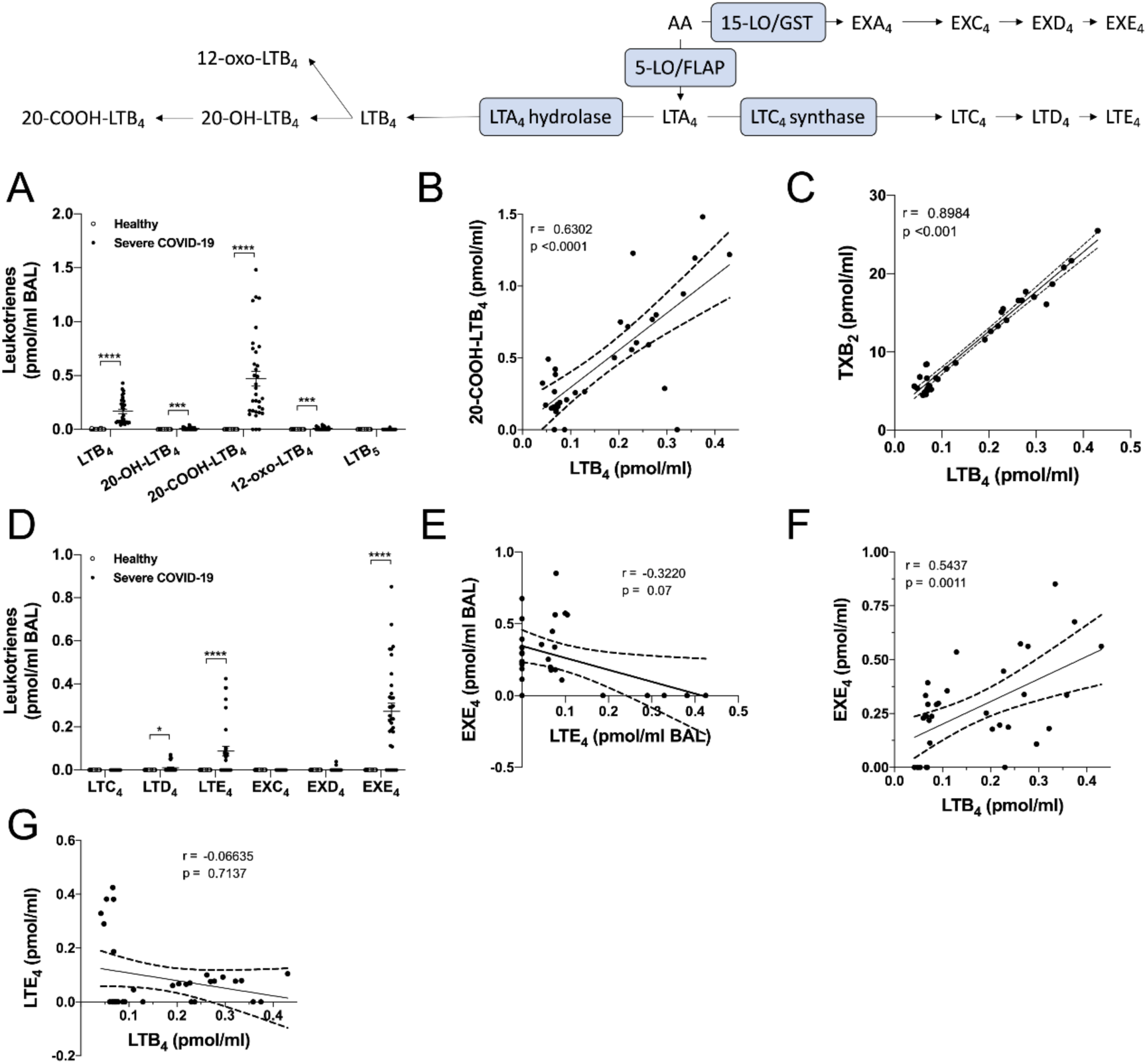
Comparison of leukotriene and eoxin levels in the BAL fluids of healthy subjects and severe COVID-19 patients. **A,D)** BALs were obtained and processed as described in methods. Lipids then were extracted and quantitated by LC-MS/MS. Results are from the BALs from 25 healthy subjects and 33 severe COVID19 patients. P values were obtained by performing a Mann-Whitney test: * p < 0.05; *** p < 0.001; **** p < 0.0001 **B,C,E-G)** Correlation tests were performed with severe COVID-19 patients by using the non-parametric Spearman’s rank. P<0.05 was the threshold of significance. The linear regression is represented with a solid line and the best-fit lines of 95% confidence are represented with dotted lines.

The prostaglandins and leukotrienes data (**figures 1 and 2**) support the idea that an increase in both arachidonic acid (AA) release and metabolism occurs in severe COVID-19 patients. As such, we investigated the levels of some fatty acids, which are the precursors of oxidized linoleic acid (LA) mediators (OXLAMs), eicosanoids and docosanoids. To that end, the levels of LA, AA, docosapentaenoic acid n-3 (DPAn-3) and docosahexaenoic acid (DHA) were all significantly increased in the BALs of severe COVID-19 patients. In contrast, the levels of eicosapentaenoic acid (EPA) did not change (**figure 3**). Of note, all tested BAL fatty acids negatively correlated with AST and ALT but positively correlated with the circulating platelet-to-lymphocyte ratio (**figure 7**).

**Figure 3.**
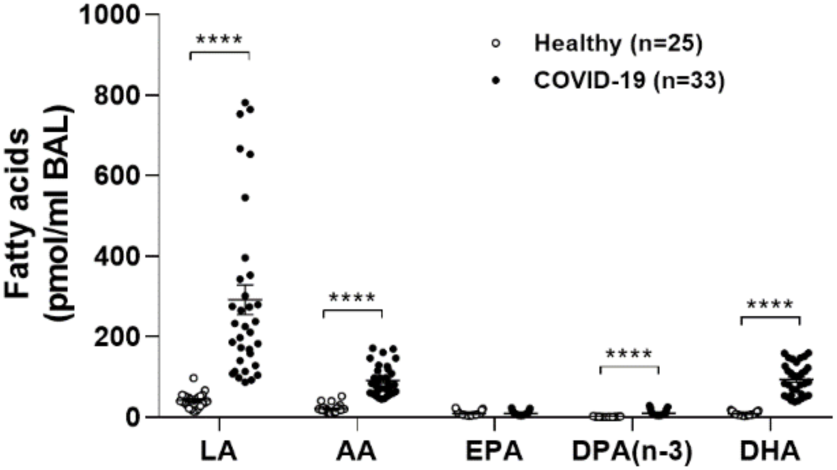
Fatty acid levels in the BAL fluids of healthy subjects and severe COVID-19 patients. BALs were obtained and processed as described in *Methods*. Lipids then were extracted and quantitated by LC-MS/MS. Results are from the BALs from 25 healthy subjects and 33 severe COVID19 patients. P values were obtained by performing a Mann-Whitney test: **** p < 0.0001.

The increased fatty acid levels (**figure 3**) and the increased in the 15-lipoxygenase-derived eoxins (**figure 2D**) led us to investigate if other fatty acid-related biosynthetic pathways were also enhanced in SARS-CoV-2-infected patients. To that end, we also quantitated additional AA-derived metabolites derived from the 12- and 15-lipoxygenases. The levels of 12-HETE and 15-HETE were also significantly increased in the BAL fluids of severe COVID-19 patients, although not to the same extent than the 5-lipoxygenase metabolite 5-HETE (**figure 4A**), indicating that AA metabolism via the 15-lipoxygenase (and possibly the 12-lipoxygenase) pathway is increased. In addition to AA, the 15-lipoxygenase pathway can metabolize numerous polyunsaturated fatty acids having a 1*Z*,4*Z*-pentadiene motif near their omega end, notably LA, EPA, DPAn-3 and DHA. We thus also assessed the levels of their most documented 15-lipoxygenase metabolites and found significant increases in 13-HODE (derived from LA), 15-HETrE (derived from dihomo-γ-linolenic acid, DGLA), 12-, 15-, and 18-HEPE, 17-HDPA(n-3) as well as 14- and 17-HDHA (**figure 4B,C**). Dual lipoxygenases metabolites from AA were also significantly increased (**figure 4D**). Of note, the levels of 15-HETE strongly correlated with those of 13-HODE (**figure 4E**) supporting an increased 15-lipoxygenase activity in the lungs of severe COVID-19 patients. Furthermore, 15-HETE levels also correlated with those of TXB_2_ (**figure 4F**). Finally yet importantly, we also detected significant increases in 9-HODE, 8-HETE, and 11-HETE, which arise from the non-enzymatic oxidation of LA and AA, pointing to an increased oxidative bust in the lungs of severe COVID-19 patients (**figure 4A,B**).

**Figure 4.**
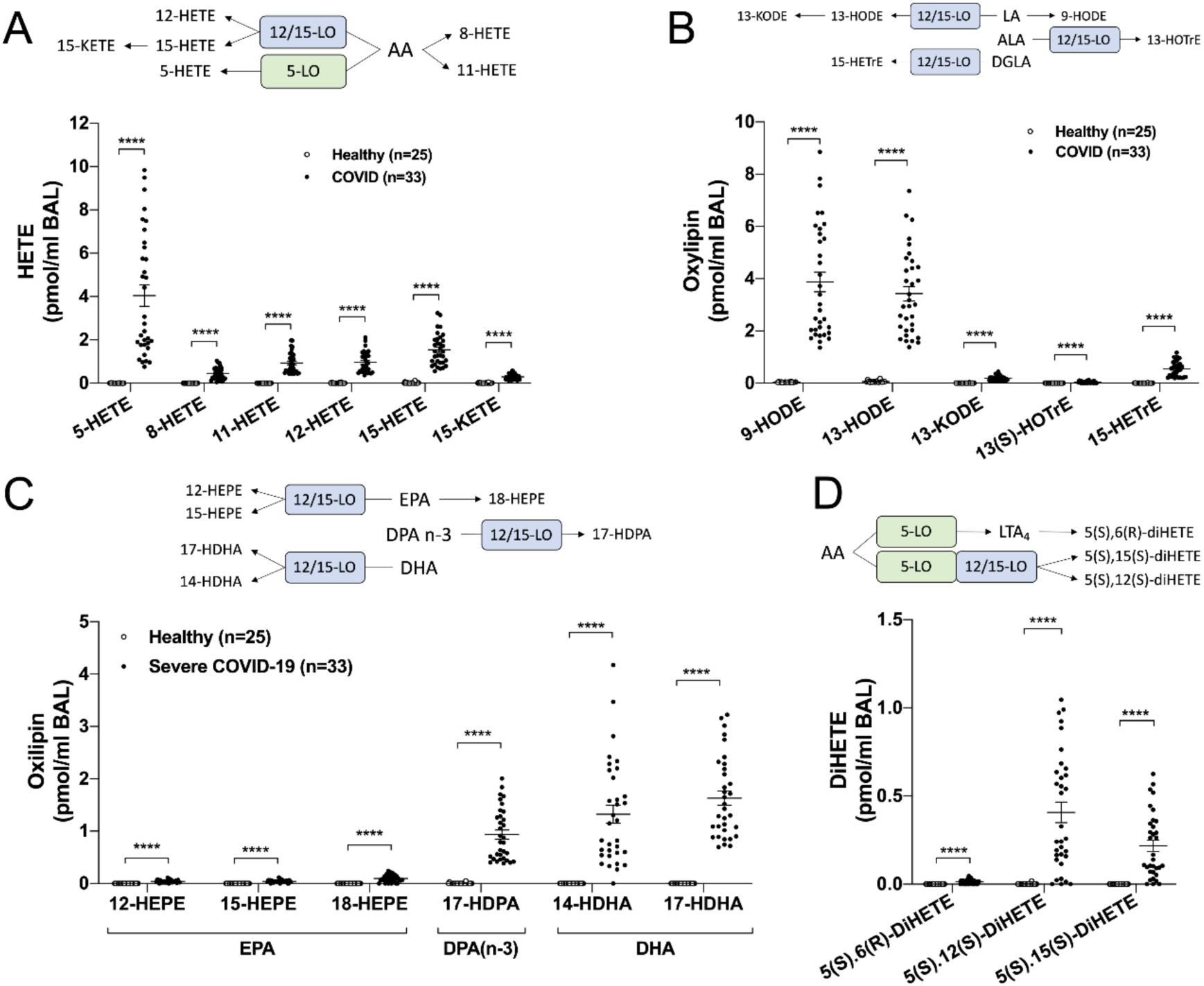
Comparison of 12- and 15-lipoxygenase-derived oxylipins and diHETEs levels in the BALs of healthy subjects and severe COVID-19 patients. **A-D)** BALs were obtained and processed as described in *Methods*. Lipids then were extracted and quantitated by LC-MS/MS. Results are from the BALs from 25 healthy subjects and 33 Severe COVID-19 patients. P values were obtained by performing a Mann-Whitney test: **** p < 0.0001.

14- and 17-HDHA, which are increased in the BALs of severe COVID-19 patients (**figure 4C**), are often regarded as good indicators of D-series resolvin and maresin levels, which are part of the ever-expanding class of specialized pro-resolving mediators (SPMs). SPMs are usually more complicated to quantitate due to their very low concentrations (10). Given their documented pro-resolving actions and anti-microbial activities, SPMs were recently postulated as being a potential approach for diminishing the inflammatory burden of COVID-19 (24). We thus investigated the levels of some commercially available SPMs. All SPMs were below detection limit in the BAL samples of healthy volunteers we analyzed. In contrast, most investigated SPMs, aside from RvD3, were detected in the BAL fluids of severe COVID-19 patients. LXA_4_ was the most prominent one, followed by RvD4, RvD5, RvD2, RVD1 and PDX (**figure 5A**). RvD5 levels correlated with those of RvD4 (**figure 5B**) but not those of DHA (**figure 5C**) indicating that the same patients are responsible for the increased D-series resolvins, which is mostly due to increased 15-lipoxygenase activity rather than an increase in DHA availability. The levels of RvD5 also correlated with those of TXB_2_ (**figure 5D)**. Again, there was no correlation between RvD5 with the clinical parameters from table 1. Unsurprisingly, the levels of LXA_4_ correlated with those of 5-HETE and 15-HETE (**figure 5E,F**), given that LXA_4_ biosynthesis requires both the 5- and the 15-lipoxygenase pathways. As for RvD5, the levels of LXA_4_ also correlated with those of TXB_2_ and the other prostaglandins (**figure 6**). Despite our good sensitivity (see table 2), we did not detect the presence or RvE1, Maresin-1 and -2.

**Figure 5.**
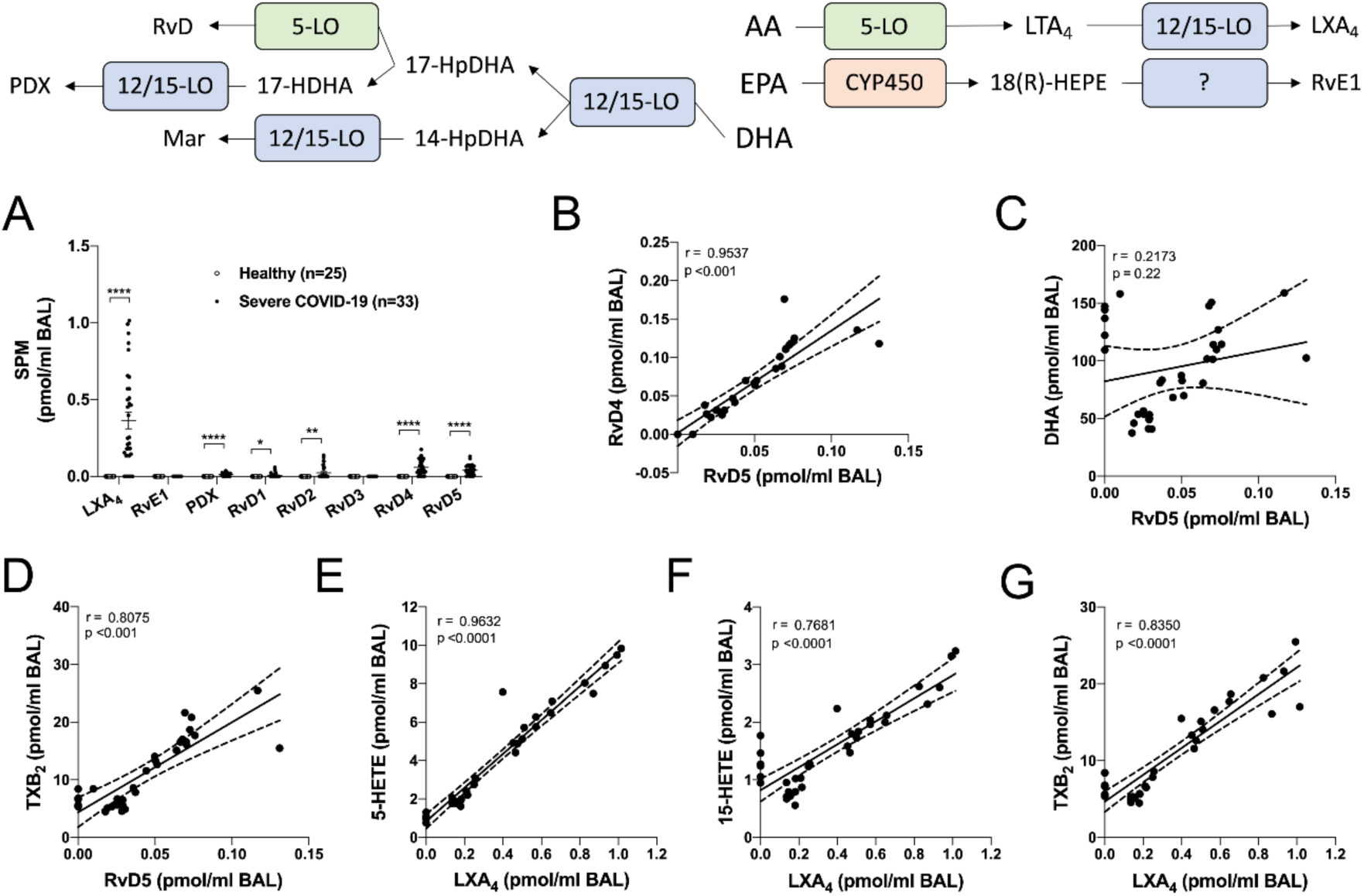
Comparison of specialized pro-resolving lipid mediator levels in the BAL fluids of healthy subjects and severe COVID19 patients. **A)** BALs were obtained and processed as described in *Methods*. Lipids then were extracted and quantitated by LC-MS/MS. Results are from the BALs from 25 healthy subjects and 33 severe COVID-19 patients.. P values were obtained by performing a Mann-Whitney test: * p < 0.05; ** p < 0.01; **** p < 0.0001. **B-G)** Correlation tests were performed with severe COVID-19 patients by using the non-parametric Spearman’s rank. P<0.05 was the threshold of significance. The linear regression is represented with a solid line and the best-fit lines of 95% confidence are represented with dotted lines.

**Figure 6.**
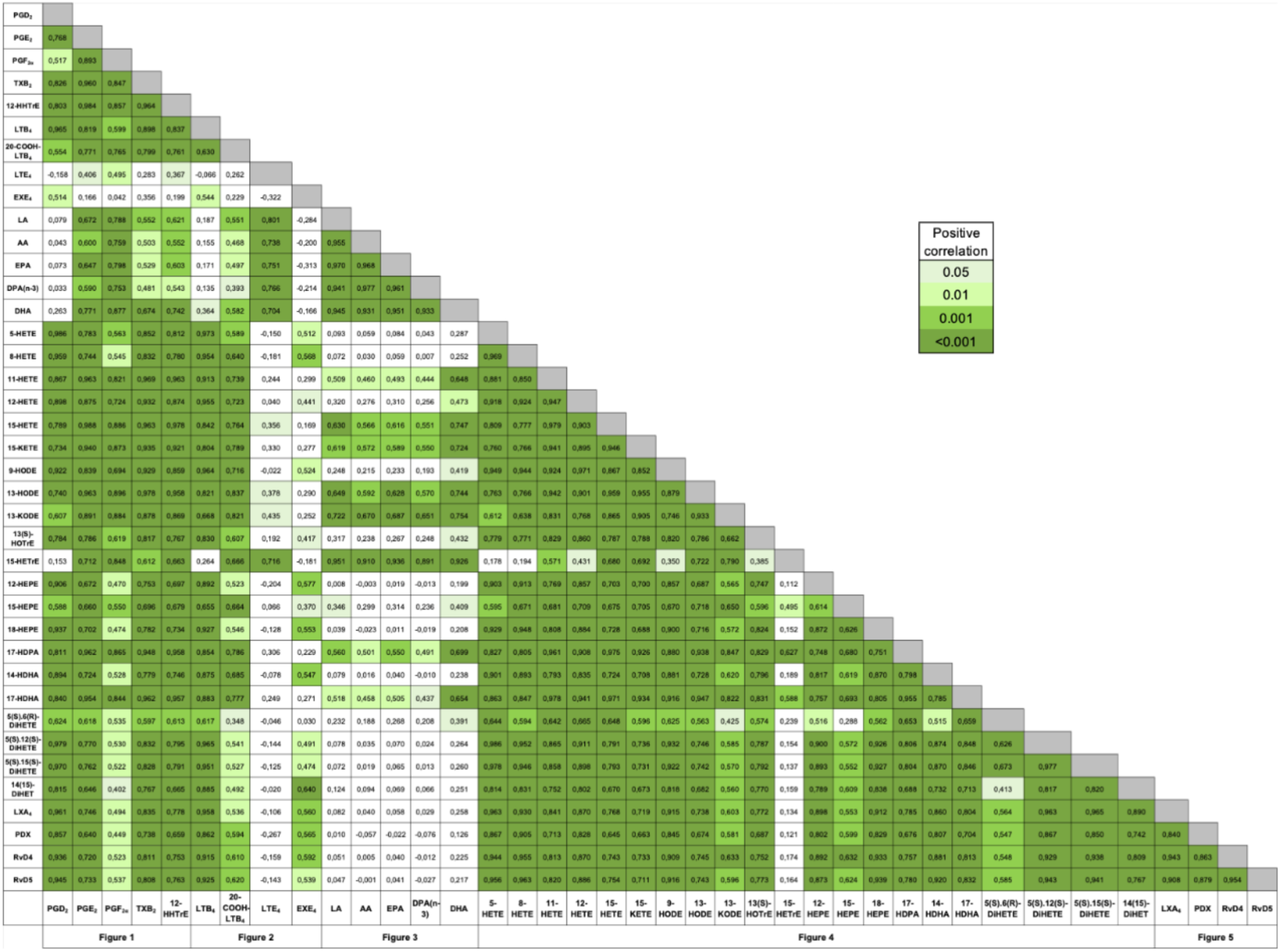
Correlation between selected bioactive lipids in the BALs of severe COVID-19 patients. Correlation tests were performed by using the non-parametric Spearman’s rank using the Graphpad Prism Software. P<0.05 was the threshold of significance. Positive correlations are shown using shades of green.

**Figure 7.**
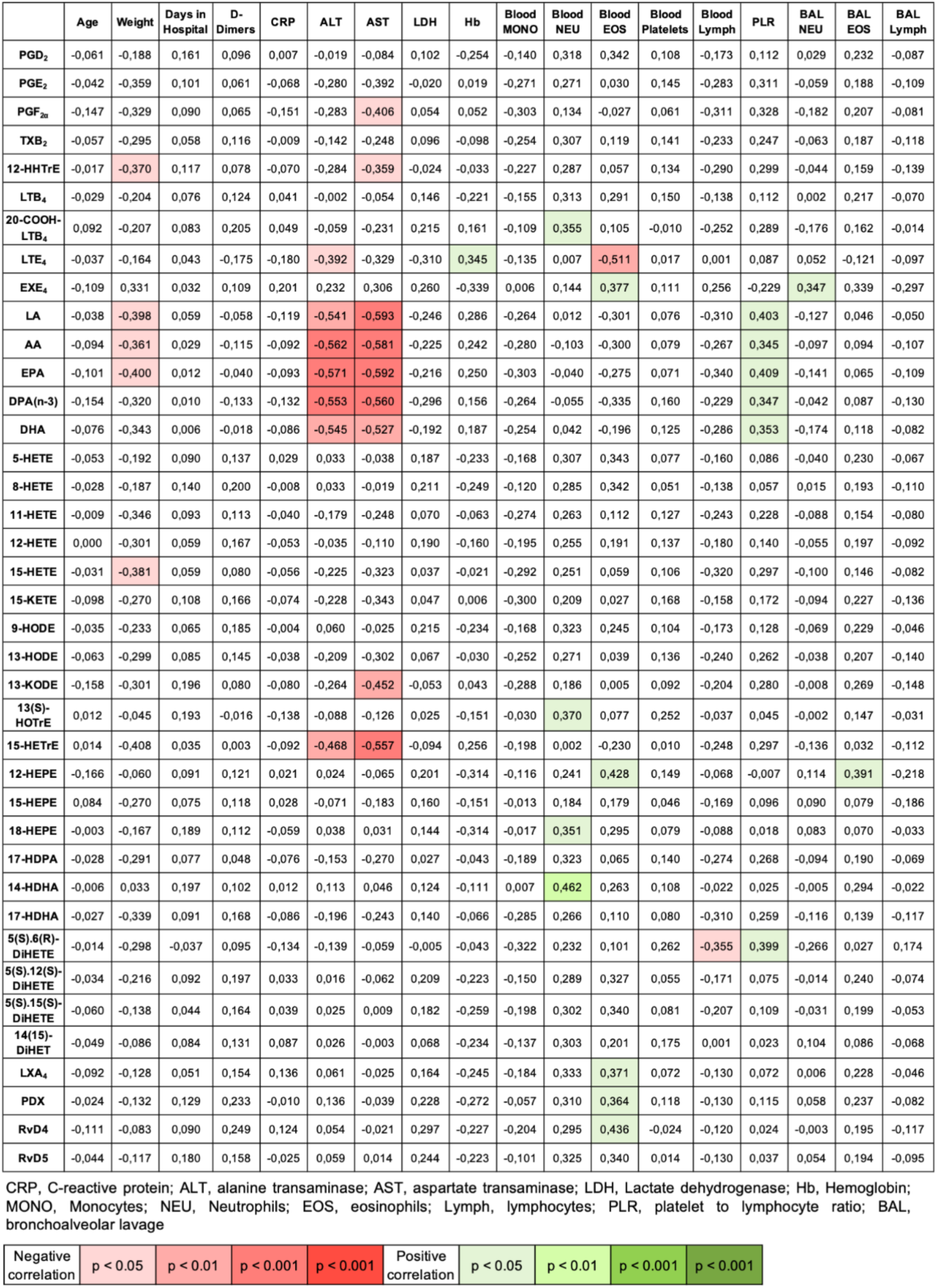
Correlation between bioactive lipids in the BALs of severe COVID-19 patients and clinical parameters. Correlation tests were performed by using the non-parametric Spearman’s rank using the Graphpad Prism Software. P<0.05 was the threshold of significance. Positive correlations are shown using shades of green and negative correlation are shown using shades of red.

When assessing the lipidome for which we could quantitate a given lipid mediator in at least 50% of severe COVID-19 patients, we can appreciate that most lipids consistently correlated with each other with few exceptions (**figure 6)**. Indeed, Fatty acids levels were not a key determinant for the levels of their metabolites, indicating that downstream enzymes (cyclooxygenases, lipoxygenases) are likely more important in that respect. Furthermore, and in agreement with their almost significant negative association (**figure 2E**), EXE_4_ and LTE_4_ correlated with different bioactive lipid subsets, LTE_4_ positively correlating with fatty acid and 15-HETrE levels while EXE_4_ mostly correlating with other 15-lipoxygenase metabolites.

## DISCUSSION

Bioactive lipids are recognized pharmaceutical targets for the treatment of numerous inflammatory diseases and lipid modulators have been utilized as such, in the last fifty years, with great success. It was thus imperative, at least to us, to define whether bioactive lipid levels were modulated in the context of severe COVID-19.

Herein, we provide clear evidences that the BALs of severe COVID-19 patients contain large amounts of bioactive lipids, which likely participate in the deleterious inflammatory response found in severe cases. More specifically, our data indicate that the BALs of severe COVID-19 patients are characterized by **1)** an increase in COX metabolites, notably the TXA_2_ metabolite TXB_2_; **2)** an increase in LTB_4_ and its metabolites; **3)** an increase in CysLT metabolites; **4)** an increase of 15-lipoxygenases metabolites derived from LA, AA, EPA, DPA and DHA; **5)** an increase in SPMs derived from AA and DHA; and **6)** a limited number of correlations between BAL lipids *vs*. blood markers and/or clinical parameters.

Cyclooxygenase metabolites were heavily increased in the BALs of severe COVID-19 patients (**figure 1**). This was somewhat expected, notably because of the increased levels of IL-1β described before (16), the latter being a good COX-2 inducer. In the lungs, COX metabolites play multiple roles. TXA_2_ promotes bronchoconstriction and activate inflammatory cells and platelets, which might be consistent with the reported enhanced platelet activation and increased thrombosis in COVID-19 (14, 25, 26). On the other hand, PGD_2_ acting *via* the DP_2_ receptor promotes the recruitment and activation of eosinophils, basophils, mastocytes and innate lymphoid cells (27-30). In contrast, PGE_2_ is usually regarded as anti-inflammatory in the lungs, notably by downregulating the pro-inflammatory functions of myeloid leukocytes such as neutrophils and macrophages, notably by activating the EP_2_ and EP_4_ receptors, ultimately leading to increased cyclic AMP levels (31). Therapies targeting the prostaglandin pathway are numerous and usually effective at controlling inflammation. Dexamethasone, a steroid improving COVID-19 symptoms and diminishing mortality (32-34), limits COX-2 expression (35-37) and might thus improve COVID-19 severity, at least in part, by diminishing the biosynthesis of prostaglandins and possibly TXA_2_. Furthermore, nonsteroidal anti-inflammatory drugs (NSAIDs) such as aspirin and ibuprofen are recognized inhibitor of cyclooxygenase activity. Early at the beginning of the pandemic, it was postulated that NSAIDs could worsen the severity of COVID-19 (38). However, a Danish study indicated that ibuprofen was not linked to severe adverse outcome (39). Furthermore, aspirin was recently shown to significantly diminish ICU admissions and the need of mechanical ventilation in an observational, retrospective study (40). Altogether, our data, combined with the recent studies on their safety in a context of COVID-19, support the hypothesis that dexamethasone and NSAIDs might diminish the inflammatory status of severe COVID-19 patients by diminishing the prostaglandins/thromboxane storm in the lungs. While TXA_2_ and PGD_2_ might have deleterious effects in the lungs, PGE_2_ and PGI_2_ might have the opposite effect, notably by inhibiting leukocyte functions (PGE_2_) and/or promoting the vasodilation of the microvasculature (PGI_2_). To that end, testing whether blocking the deleterious effects of PGD_2_ and TXA_2_ with the dual DP_2_/TP antagonist Ramotraban might be beneficial in this particular context, as proposed recently (41).

Prostaglandins and TXA_2_, as assessed by quantitating TXB_2_ levels, were not the only eicosanoids that were increased in the BALs of severe COVID-19 patients. Indeed, leukotrienes were also increased, notably LTB_4_, LTE_4_, and EXE_4_ (**figure 2**). This indicates that in addition of the deleterious effects that some COX metabolites might have, those linked to LTB_4_ and CysLTs might also impair lung functions and participate in the inflammatory burden in the lungs of severe COVID-19 patients (22). While LTB_4_ has previously been shown to stimulate host defense (41), notably during viral infections, the ω-LTB_4_/LTB_4_ ratio of 2.84 we observed is more reminiscent of what is found in cystic fibrosis and severely burned patients, two conditions in which infections are not well controlled (42-45). Conceptually, this might diminish the effectiveness of LTB_4_ at promoting its host-defense related functions, as we recently published (46). Interestingly, there was a trend toward an inverse correlation between LTE_4_ and EXE_4,_ indicating that these cysteinyl-LTs are likely coming from different cellular sources. This was supported by the differential and significant correlations between the different lipids we observed, LTE_4_ and EXE_4_ differentially correlating with distinct lipids (**figure 6**). To that end, EXE_4_ levels in the BALs positively and significantly correlated with blood eosinophils counts. While there was a strong trend between EXE_4_ and BAL eosinophils counts the latter correlation had a p value of 0.0536. Altogether, this suggests that the presence and/or biosynthesis of eoxins is, in part, linked to eosinophils. A therapeutic approach to target LTs from the 5-lipoxygenase pathway could be the use of Zileuton, which is commercially available for the management of asthma and has a good safety profile (47).

Another pharmaceutical approach that could simultaneously diminish the levels of COX and 5-lipoxygenase-derived eicosanoids could be the use of phosphodiesterase 4 inhibitors such as Roflumilast. While there is no evidence of phosphodiesterase 4 inhibitors’ efficacy in the management of SARS-CoV-2-infected people (48), they might diminish inflammation by preventing the degradation of cyclic AMP, increasing intracellular cAMP concentrations and decreasing the release of AA, the precursor of prostaglandins and leukotrienes. Furthermore, cAMP-elevating agents such as adenosine and PGE_2_ are recognized anti-inflammatory autacoids that inhibit numerous leukocyte functions such as eicosanoid biosynthesis, oxidative burst, calcium mobilization and migration (49-53). While phosphodiesterase 4 inhibitors might diminish the levels of PGE_2_ by diminishing AA levels, they would nonetheless amplify the inhibitory effects of the remaining PGE_2_, as recently demonstrated *in cellulo* (54).

While it is largely accepted that the group IVA cytosolic phospholipase A_2_ mediates most of AA release in activated human leukocytes (e.g. in (55)), the increased levels of LA, EPA and DHA support the idea that other phospholipases A_2_ are also heavily involved in the lipidome we have quantitated. Those additional phospholipases likely belong to the superfamily of secreted phospholipases A_2_, which can hydrolyze phospholipids containing fatty acids other than AA (56, 57) and are reported to contribute to lung inflammation (58-61). However, and assuming that a large part of eicosanoids is the consequence of the group IVA cytosolic phospholipase A_2,_ our findings do not allow us to pinpoint which other phospholipases are involved in the release of LA, AA, EPA, DPA and DHA.

The metabolism of EPA, DPA and DHA, which are omega-3 fatty acids, often results in the biosynthesis of SPMs, most of which are anti-inflammatory and pro-resolving lipids. Their presence in high levels was somewhat surprising as they are generally thought to appear during the resolution of inflammation, after the biosynthesis of prostaglandins and leukotrienes has occurred and following a class-switch from pro-inflammatory to pro-resolving lipids (62). However, our data indicate that prostaglandins, leukotrienes and SPMs can certainly co-exist and participate in the inflammatory cascade during the acute phase of inflammation/infection in which resolution has not been fully engaged. Moreover, given their strong documented anti-inflammatory effects and their host-defense boosting functions, they could be viewed as potential mediators to enhance to limit the infection. To that end, it was recently proposed that dexamethasone could participate in the upregulation of SPM biosynthesis and effects in moderate to severe COVID-19 patients, although this needs to be confirmed (63). While omega-3 fatty acids, notably purified preparation enriched in EPA and DHA are commercially available as dietary supplements, we did not see a correlation between SPMs and the levels of EPA or DHA, indicating that SPMs levels are likely more dependent on biosynthetic enzymes other than phospholipases such as lipoxygenases than EPA or DHA availability. As of today, no resolution pharmacology treatment has been approved by governmental health agencies yet. However, some putative E- and D-series resolvin precursors, notably 18-HEPE and 17-HDHA, are available as dietary supplements.

In addition to cyclooxygenase and lipoxygenase derivatives, other bioactive lipids are recognized, at least in mice, to downregulate the immune response. This is notably the case of the endocannabinoids *N*-arachidonoyl-ethanolamine (AEA) and 2-arachidonoyl-glycerol (2-AG), which are AA-derived lipids (64). The anti-inflammatory effects of AEA and 2-AG are mostly related to the activation of the CB_2_ receptor, which is heavily expressed by leukocytes (65). In addition to 2-AG and AEA, their congeners from the monacyl-glycerols and *N*-acyl-ethanolamines and *N*-acyl-aminoacids families, now viewed as the endocannabinoidome, could potentially regulate the inflammatory response during SARS-CoV-2 infections, notably by activating other anti-inflammatory receptors (66). It will thus be crucial to decipher whether the endocannabinoidome is modulated during COVID-19 and if a potential target such as CB_2_ receptor agonists, or endocannabinoid hydrolase inhibitors could be helpful in the treatment of COVID-19 or other coronavirus-mediated infections.

Nonetheless the expanded definition of a dysregulated lipidome we unravel in SARS-CoV-2 infections requiring mechanical ventilation, our study has limitations. **1)** BALs from healthy controls are from younger individuals than those from severe COVID-19 patients. This could have an impact on some mediators, notably PGD_2_ levels, which are increased in aged mice and worsen SARS-CoV infections (67); **2)** The procedure to obtain BAL fluids is slightly different between healthy controls and severe COVID-19 patients. Indeed, while the volume of saline injected to healthy people was 50 ml, that of severe COVID-19 patients was 100 ml. The possible outcome of this discrepancy could be and underestimation of lipid levels in one group vs. the other. While some might argue that the first saline bolus contains more lipids than the following one(s), other might argue that lipid levels are consistent in each bolus. Given that, in our hands, the number of BAL cells is usually greater in the first bolus than in the subsequent ones (when multiple boli are utilized), we are confident that the BAL fluid from the first bolus likely contains more lipids than the second one. This would thus lead to a slight underestimation of lipids in the COVID-19 group. **3)** Unfortunately, the leukocyte counts in the BALs of severe COVID-19 patients did not include the number of alveolar macrophages and it is thus impossible to determine a clear differential count of leukocytes in those samples. Furthermore, the eosinophil counts were somewhat limited by the apparatus as it could estimate the counts to +/- 10^5^/ml cells, which likely has an impact on the different correlations involving eosinophils, notably with EXE_4_; **4)** We could not compare the levels of lipids in the BALs of severe COVID-19 patients to those found in the blood. This appears important given the limited number of correlations found between BAL lipids and other circulating markers such as CRP and D-dimers. To that end, a recent study indicated differences in some of the herein investigated bioactive lipids in the circulation (68). It will thus be important to pursue our investigations and determine whether there is a transposition of the BAL lipidome to the circulation.

In conclusion, our data unmask that in addition to the recognized cytokine storm previously documented, lung fluids of SARS-CoV-2-infected patients indicate that a lipid storm also occurs. As highlighted above, the rich library of governmental health agencies-approved lipid modulators might provide beneficial and usually low-cost add-ons to the current therapeutic arsenal utilized to diminish the severity of COVID-19 and possibly additional coronavirus-mediated infections.

## Data Availability

All relevant data are within the paper and its Supporting Information files.

## ACKNOWLEDGEMENTS

ASA, AC, VR, ML, and NF are members of the Quebec Respiratory Health research Network.

